# External Validation of Machine Learning Models for Traumatic Brain Injury: Performance, Efficiency, and Carbon Imprint

**DOI:** 10.64898/2026.07.05.26357337

**Authors:** Tobias Gauss, Théotime Fehr Delude, Alexandre Kalimouttou, Oussama Seddiki, Caroline Sanchez, Jules Grèze, Clément Brossard, Jean-Denis Moyer, Geoffray Brelurut, Sofiane Medjkoune, Alexandre Krainik, Thomas Boulier, Kevin Lagarde, Arnaud Lazard, Pierre Bouzat, Benjamin Lemasson

## Abstract

**Background:** Machine learning (ML) models for traumatic brain injury (TBI) prediction increasingly demand extensive data, computational resources, and energy consumption, yet simpler models may offer comparable clinical benefit with lower barriers to deployment. This study compares predictive performance, computational efficiency, carbon footprint, and real-world feasibility of resource-efficient (“pauci-parameter”) versus data-intensive (“multiparameter”) ML models for predicting TBI patient care pathways and outcomes.

**Methods:** External validation study in a level 1 trauma center (n=534 adult TBI patients with GCS<9 and/or intracranial injuries). Seven models tested: two pauci-parameter models using only routine prehospital variables (PREHOSP, 15 variables) or CT image analysis (CT-TIQUA), and five multiparameter models integrating clinical and imaging data. Primary outcome: positive likelihood ratio for predicting neurocritical care intensity, mortality (7/30-day, 6-month), and functional outcome (Glasgow Outcome Scale Extended). Secondary outcomes: computation time, carbon footprint, clinical implementability.

**Results:** Multiparameter models showed superior performance but did not consistently translate to better clinical utility. PREHOSP (pauci-parameter) showed comparable performance to complex models for most outcomes. The best-performing multiparameter model (MULTI-PRE) required 100-fold longer inference time and 10-fold higher carbon emissions per prediction versus simple models, while net clinical benefit was nearly identical (0.06 vs 0.05). Models using only prehospital data demonstrated greater generalizability and lower deployment barriers.

Interpretation

Computational complexity and resource intensity should factor equally with predictive performance in clinical AI deployment decisions. For sustainable digital health implementation—especially in resource-limited settings—simpler models with comparable clinical benefit may enable broader access while reducing environmental and financial costs.

**Funding:** Fondation Gueules Cassées, Grant 27-2023

**AUTHOR SUMMARY:** *Why Was This Study Done?:* Artificial intelligence (AI) models are increasingly used in hospitals to help doctors predict which brain injury patients need intensive care and what their outcomes might be. However, many published models are very complex. They use multiple variables and require expensive computer systems to run. Hospitals often struggle to implement these complex models because they require operationalisation barriers.

– Multiple data systems that don’t easily communicate
– Expensive computer infrastructure
– Highly trained technical staff
– Energy-intensive processing

This raises the question whether complex models actually work better than simpler ones? If simple models work just as well, they could be used in many more hospitals especially in lower-resource countries where complex systems aren’t available.

*What Did the Researchers Do?:* The team compared seven different Machine and Deep Learning models for predicting brain injury patient needs and outcomes in 534 patients treated at a major trauma center in France. They tested:

– Two simple models using only routine prehospital data (vital signs, Glasgow Coma Scale)
– Five complex models using 40+ variables from multiple hospital computer systems plus CT scan segmentation analysis

They compared the models’ accuracy, how long they took to run, how much electricity they used (carbon footprint), and how easy they would be to implement in different types of hospitals.

*What Did They Find?:* The simple models worked almost as well as the complex ones. For example:

– Simple model (PREHOSP): Predicted mortality with reasonable accuracy
– Complex model (MULTI-PRE): Predicted mortality slightly better, but required 100 times more processing time and 10 times more electricity

When measured by “clinical benefit”, the number of correct treatment decisions made per 100 patients, both simple and complex models performed similarly (0.05 vs 0.06 additional correct decisions). The complex model’s training generated as much greenhouse gas as a clinical CT scan. For hospitals committed to environmental sustainability, this matters.

*Why Is This Important?:* This study challenges the common assumption that more complex models perform better. The findings have practical implications: For well-resourced hospitals: Complex models may offer only modest benefit that may not justify their cost and complexity. For hospitals with limited budgets: Simple models may provide comparable accuracy without expensive infrastructure. For low-income countries: Simple models using prehospital data may be deployed immediately in emergency medical services, even without integrated hospital computer systems. For environmental sustainability: Simpler AI models consume far less energy, an important as hospitals worldwide commit to net-zero emissions and will implement energy intense decision support tools. What Are the Clinical Implications? The authors recommend:

1. Asses clinical performance, operational barriers and carbon imprint when comparing simple and complex models before implementation and deployment of decision support tools
2. Reserve complex models for specialized high-resource trauma centers where infrastructure supports them
3. Validate simple models in diverse care settings (ambulance services, regional hospitals, low-income countries) to confirm they work everywhere

*What Comes Next?:* The next step is testing the simple model in real-world settings across different countries and hospital types to prove it works reliably everywhere.

## INTRODUCTION

Outcome prediction in neurotrauma remains challenging even for experienced clinicians and is affected by cognitive bias (1) Machine learning algorithms theoretically provide automated, consistent decision support to avoid these limitations. However, most prediction models target single outputs such as intracranial pressure (ICP), mortality, or long-term outcome,(2,3) with few anticipating required neurocritical care levels and none addressing the entire traumatic brain injury (TBI) patient pathway.

Pauci-parameter models rely on limited variables from single data sources, while multiparameter models integrate diverse predictors from clinical variables, biomarkers, radiomics, and intensive care unit data.(4) Although multiparameter approaches theoretically offer informational gain, they require systems capable of accessing and processing data from multiple fragmented sources. The lack of standardized formats impairs real-time data access, making information retrieval, integration, and deployment resource-intensive without proven clinical benefit.(5,6) In contrast, pauci-parameter models use routinely available inputs and may offer comparable performance with lower resource consumption and lower carbon imprint.(7)

This question has strategic importance as health institutions will increase deployment of AI tools, these tools will consume server capacity, drive energy consumption, challenge the hospital informatic infrastructures and increase carbon footprint.(8) Less resource-intensive algorithms could be easier to deploy with acceptable performance lower energy consumption and lower carbon footprint. This trade off between predictive performance and computational and energetic and ecological efficiency remains a knowledge gap. This gap challenges operationality and implementation of enhanced decision tools in critical care.(5,9)

To investigate this gap, we capitalized on two previously developed prediction models (PREHOSP and CT-TIQUA) to perform this comparative study. Both models predict actionable patient needs along the TBI pathway. PREHOSP relies exclusively on routine prehospital predictors to predict neurosurgery and ICP monitoring using machine learning.(10) CT-TIQUA employs a radiomic approach of admission CT scans to predict neurocritical care levels.(11)

The primary objective was external validation of PREHOSP and CT-TIQUA. The secondary objective compared performance and calculation time, cost and carbon footprint of these two pauci-parameter models with five multiparameter models integrating all available prehospital and resuscitation room variables. We hypothesized that multiparameter models would provide better prediction of the TBI patient pathway than pauci-parameter models at slightly higher cost and with an acceptable carbon footprint.

## METHODS

### Study Design and Setting

This retrospective single-center comparative and external validation study was conducted at Grenoble University Hospital (GUH), a level 1 trauma center with dedicated neurocritical care. Patients were managed according to standard operating procedures based on national and international guidelines. The institutional review board of Grenoble University hospital approved the protocol on the 22/11/2024 (File nr 38RC23.0351, CHU Grenoble, Grenoble, France). All patients or their next of kin received written and formal notification with the opt-out possibility to opt-out of the study; four patients decided to opt out. The manuscript follows AI APPRAISE TOOL guidelines (12).

### Patient Selection

All adults admitted to the trauma resuscitation unit between August 1, 2020, and December 31, 2021, were screened. Inclusion criteria comprised TBI defined as Glasgow Coma Scale <9 and/or documented intracranial injuries on admission CT scan. Data were retrieved by two expert clinicians blinded to model outputs. CT scans were retrieved from hospital imaging systems and stored anonymously. Observations with missing predictor variables or outcomes were excluded without imputation. The corresponding CT scan was retrieved from the hospital image processing system and corresponding ID assigned. The anonymized CT scan with corresponding ID was stored on a safe imaging platform (*shanoir*) in DICOM format. All data (clinical and images) were stored on a safe server after anonymization and ID number assignment and data management performed centrally by professionals blinded to the model output. Figure 1 summarizes the flowchart and data processing.

**Figure 1.**
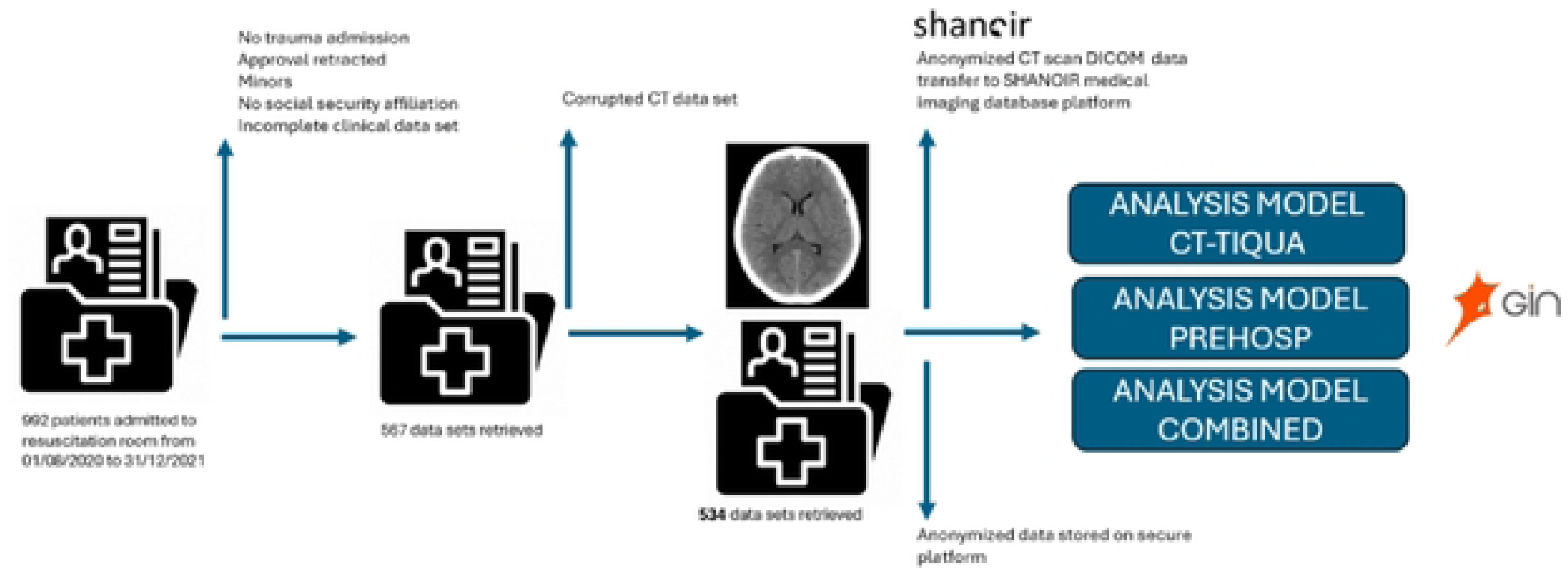
**Study data flow**

Prediction Models

### Pauci-Parameter Models

PREHOSP uses 15 routinely collected prehospital predictors (Glasgow Coma Scale components, age, capillary haemoglobin, blood pressure, heart rate, pupil anomaly, limb ischemia, pelvic injury, cardiac arrest, external bleeding, penetrating trauma, vasopressor use). This classification model employs light Gradient Boosting,(13) initially trained to predict emergency neurosurgery within 24 hours and need for ICP monitoring (supporting material 2).

CT-TIQUA analyses admission CT scans performed within 60 minutes of resuscitation room admission. The model performs four steps: extraction (removes bone), registration (locates lesions topographically), segmentation (identifies injury types using deep learning), and quantification (computes volumes). CT-TIQUA recognizes seven distinct TBI classes using Convolutional Neural Networks: intraparenchymal, subdural, epidural hematoma, intraventricular and subarachnoid hemorrhage, petechiae, and edema (supporting information 3). The initial CT-TIQUA model was improved, first by testing three different brain extraction methods; the TOTALSEGMENTATOR (14) approach proved to be the best performing with an AUC of 0.98 tested against manual ground truth brain masks. Second, three different *atlas registration* techniques were explored; a combination of Histogram matching and Advanced Normalization Tools appeared to be the most reliable.(15) The supporting information provides details on the development process.

### Multiparameter Models

Five multiparameter models were developed: MULTI (combining PREHOSP and CT-TIQUA features), PREHOSP-X (relying on all available 23 prehospital variables), RESUS-X (relying on all available 18 resuscitation room variables), MULTI-PRE (PREHOSP-X combined with CT-TIQUA), and MULTI-RESUS (RESUS-X combined with CT-TIQUA). PREHOSP-X and RESUS-X employed light Gradient Boosting (supporting information 4).

In summary, seven models were available. Two pauci-parameter models PREHOSP and CT-TIQUA and five multi-parameter models, MULTI, PREHOSP-X, RESUS-X and MULTI-PRE and MULTI-RESUS.

### Model Outputs

All seven models predicted (Figure 2):

1. Highest neurocritical care intensity within seven days per TILSUM classification (dichotomized as >level 0); http://www.tbi-impact.org/cde/mod_templates/T_TIL.9.1.pdf)(16)
2. Highest neurocritical care per Seattle International Severe TBI Consensus Conference (SIBICC)(17) (dichotomized as >tier 0)
3. Mortality at 7-day, 30-day, and 6-months
4. Extended Glasgow Outcome Scale at 30-day and 6-months

**Figure 2.**
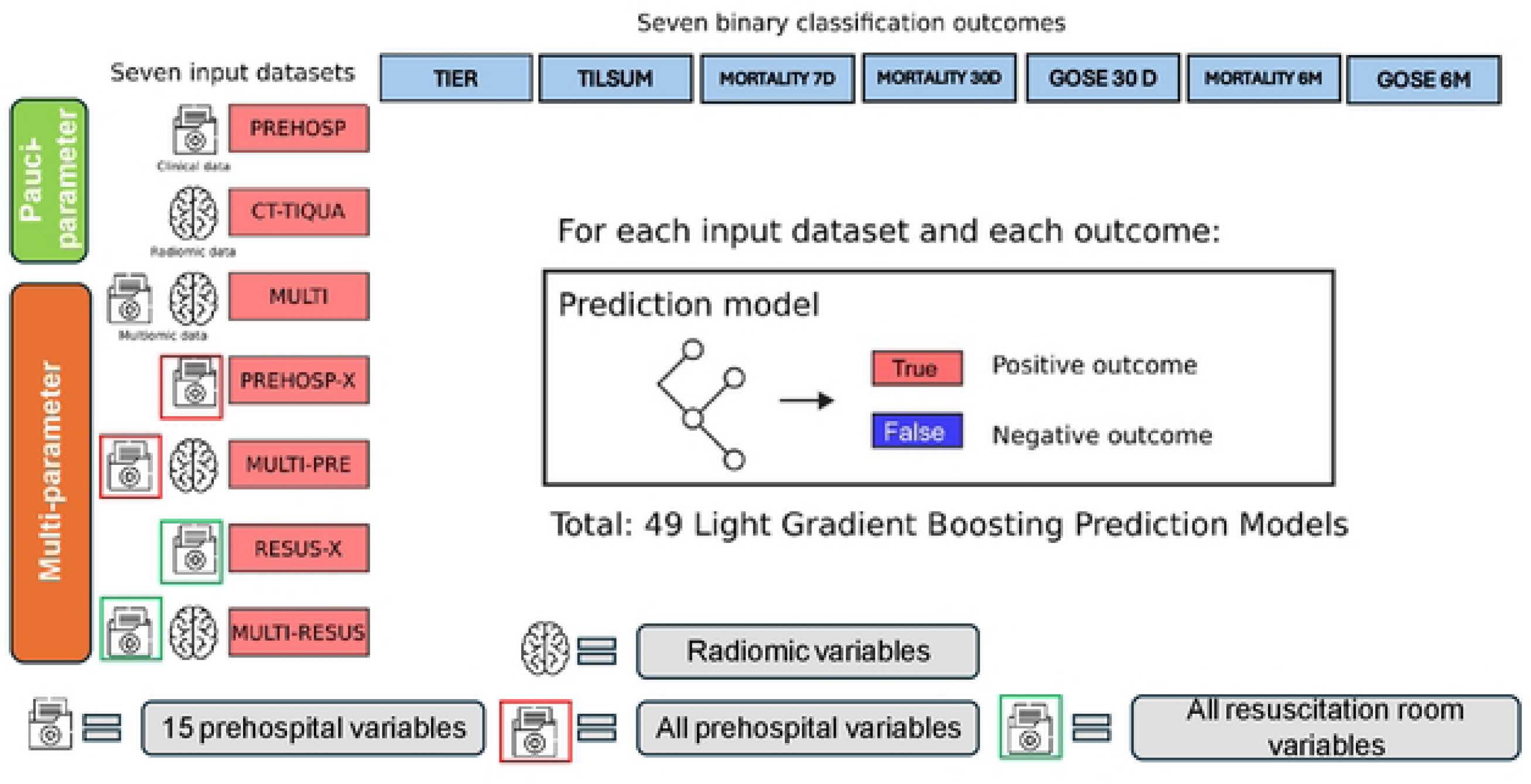
Model type and outcome prediction per type of predictors; SIBICC= Seattle International Traumatic Brain Injury Consensus Conference. TILSUM=Therapeutic Intensity Level. D= day. M= Month. GOSE= Glasgow outcome scale extended

Each prediction constituted binary classification. Models underwent hyperparameter optimization, class imbalance correction, and cross-validation (see supporting information for details).

### Primary assessment criterion

The primary assessment criterion was the positive Likelihood ratio with 95% confidence interval. All other metrics were considered as secondary outcome. The primary analysis concerned the comparison between the positive likelihood ratios between the pauci-parameter models PREHOSP and CT-TIQUA and all multi-parameter models (MULTI, PREHOSP-X, RESUS-X and MULTI-PRE and MULTI-RESUS) for all seven outputs (see above). All other metrics, execution time and the additional analysis recruiting all available prehospital and resuscitation room variables (preceding paragraph) were considered as secondary outcome criteria.

### Sample size calculation

The sample size was determined based on the comparison of the light Gradient Boosting prediction and the truly observed rate of the composite outcome criterion *need for neurosurgery and ICP probe placement*. The prevalence of the outcome in a pilot study was 15%, sensitivity 84% and specificity of 87% for the light Gradient Boosting based on a pilot study.(10) With a type 1 error of 5%, a prevalence of the main output of 15% and to obtain a positive likelihood ratio around 6 for 95% confidence interval with a width of 0.255, a sample of 550 patients was necessary. (18)

### Computation efficiency and Carbon Footprint calculation

Computation time for training and single case inference for the Machine Learning and Deep learning (radiomics) models were expressed in seconds. The respective carbon footprint were calculated from development to application, The method used has been described at https://codecarbon.io/. In summary, the following formula is used:

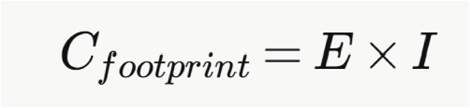

Where *Cfootprint* is carbon footprint in kg of CO₂ equivalent (CO₂eq), *E*= energy consumed by the computation in kilowatt-hours (kWh), *I*= carbon intensity of electricity in the geographic location of the hardware, in kg CO₂eq per kWh. CodeCarbon computes the carbon footprint of your code by measuring at fixed intervals, e.g. every 15 seconds, the electricity consumption of the GPU, CPU and RAM on which the code is executed. The package also monitors the duration of code execution and uses this information to compute the total electricity consumption. CodeCarbon retrieves information about the carbon intensity of the electricity in the geographic location of the hardware. The calculation of the carbon footprint included the development and training phase of each model.

### Data analysis

All data were analyzed by a data engineer and clinician researcher (TFD, AK) supervised by a senior clinician researcher (TG) and two research directors (BL, JJ). Continuous data were described as median (quartiles 1–3) and categorical variables as count (percentages).

The true observed rate of all listed outputs in the patient cohort was considered as reference. The individual (pauci-parameter) and combined (multi-parameter) model performance was assessed using the following metrics: AUC ROC, sensitivity, specificity, positive, negative predictive value, Brier score, Youden Index, F2, accuracy and precision and the positive and negative likelihood ratio (LR+ and LR-). The positive likelihood ratio of the seven models considered for each model and their combination were compared through a z-score with a 95% confidence interval computed by bootstrapping, estimate the standard error from the CI and apply pairwise z-tests on log(LR+) values with Bonferroni correction for multiple comparisons. The net clinical benefit for each prediction model and combined use was calculated according to Vickers et al with a decision threshold of 10%. (19) Potential bias of age and sex were compared for all metrics. All calculations and developments were performed on Python (version 3.11.0) or R and R studio (version 2024.12.1).

## RESULTS

### Patient Characteristics

Of 992 patients admitted to the resuscitation room and screened, 567 met inclusion criteria and 534 had complete datasets (Figure 1). Mean age was 47.3 years (SD 22), 75% were male (400/534), and 96% (517/534) sustained blunt trauma. Median prehospital Glasgow Coma Scale was 15 [IQR 14,3], with 6% (32/534) presenting a shock index >1.1. Mean Injury Severity Score was 12.7 (SD 11.7), with 11% having Abbreviated Injury Scale head >3. ICU admission occurred in 39% (210/534), with 13% (74/534) requiring SIBICC tier >0 and 29% (144/534) requiring TILSUM >0. Mortality was 5.2% (27/534) at all time points. At 6 months, 9% (50/534) had Glasgow Outcome Scale Extended <8 (Table 1).

**Table 1.** Patient characteristics. SIBICC= Seattle International Traumatic Brain Injury Consensus Conference. TILSUM=Therapeutic Intensity Level.

| Variable | Total n=534 |
| --- | --- |
| Age (mean, SD) | 47.27 (22) |
| Sex Female (%) | 134/534 (25%) |
| Blunt/penetrating mechanism (%) | 517/17 (96.7/3.3) |
| Prehospital systolic blood pressure, mean (SD) | 123.13 (29) |
| Prehospital GCS, median [IQR] | 15.00 [14, 3] |
| Prehospital GCS motor, median [IQR] | 6 [6, 1] |
| Prehospital abnormal pupil | 30 (5.8) |
| Prehospital Intubation | 66 (12) |
| Prehospital Osmotherapy | 0.06 |
| Abbreviated Injury Scale Head > 3 (%) | 62 (11) |
| ISS, mean (SD) | 12.70 (11.7) |
| IGS, mean (SD) | 23.69 (17.5) |
| Marshall Score (%) | 1.42 |
| 1 | 427 (80) |
| 2 | 52 (9) |
| 3 | 15 (2.8) |
| 4 | 16 (3) |
| 5 | 11 (2) |
| 6 | 10 (2) |
| Intraparenchymal hematoma (%) | 55 (11) |
| Subdural hematoma (%) | 64 (12) |
| Epidural Hematoma (%) | 15 (3) |
| Intraventricular hemorrhage (%) | 25 (5) |
| Subarachnoid hemorrhage (%) | 90 (17) |
| Petechiae (%) | 50 (10) |
| Oedema (%) | 33 (6) |
| Diffuse axonal injury (%) | 13 (3) |
| Neurocritical care therapy |  |
| Admission critical care (%) | 210 (39) |
| Ventilation (%) | 120 (22) |
| Intracranial pressure probe (%) | 45 (9) |
| Extraventricular drainage (%) | 21 (4) |
| Deep sedation (%) | 29 (5.5) |
| Muscle Paralysis (%) | 15 (3) |
| Craniectomy (%) | 11 (2) |
| SIBICC > tier 0 (%) | 74 (13) |
| TILSUM > level 0 (%) | 144 (29) |
| Withdrawal of care (%) | 17 (3) |
| 7 day mortality (%) | 27 (5.2) |
| 30 day mortality (%) | 27 (5.2) |
| 6 month mortality (%) | 27 (5.2) |
| 30 day Glasgow Outcome scale extended, class < 8/7 (%) | 63 (11%) |
| 6 month Glasgow Outcome scale extended, class < 8/7 (%) | 50 (9%) |

### Primary Outcome: Likelihood Ratios

Multiparameter models were not systematically superior to pauci-parameter models. Positive likelihood ratios ranged from 3.8 (95% CI 2.57-5.47) for CT-TIQUA predicting TILSUM to 41.83 (95% CI 21.08-78.95) for MULTI-PRE predicting 6-month Glasgow Outcome Scale Extended. Models demonstrated higher likelihood ratios for mortality and neurological outcome than neurocritical care level predictions. Likelihood ratios for TILSUM and SIBICC did not exceed 10, while all models achieved ratios above 10 for mortality and outcome predictions.

PREHOSP showed no significant differences in positive likelihood ratio compared with multiparameter models for TILSUM, SIBICC, 30-day and 6-month Glasgow Outcome Scale Extended. PREHOSP had significantly lower positive likelihood ratios than MULTI-PRE for 7-day (13.82, 95% CI 8.79-19.44 vs 40.30, 95% CI 23.26-75.93), 30-day (14.68, 95% CI 9.21-21.81 vs 38.70, 95% CI 21.93-73.79), and 6-month mortality (14.19, 95% CI 9.11-21.10 vs 41.83, 95% CI 21.08-78.95; see figure 3).

**Figure 3.**
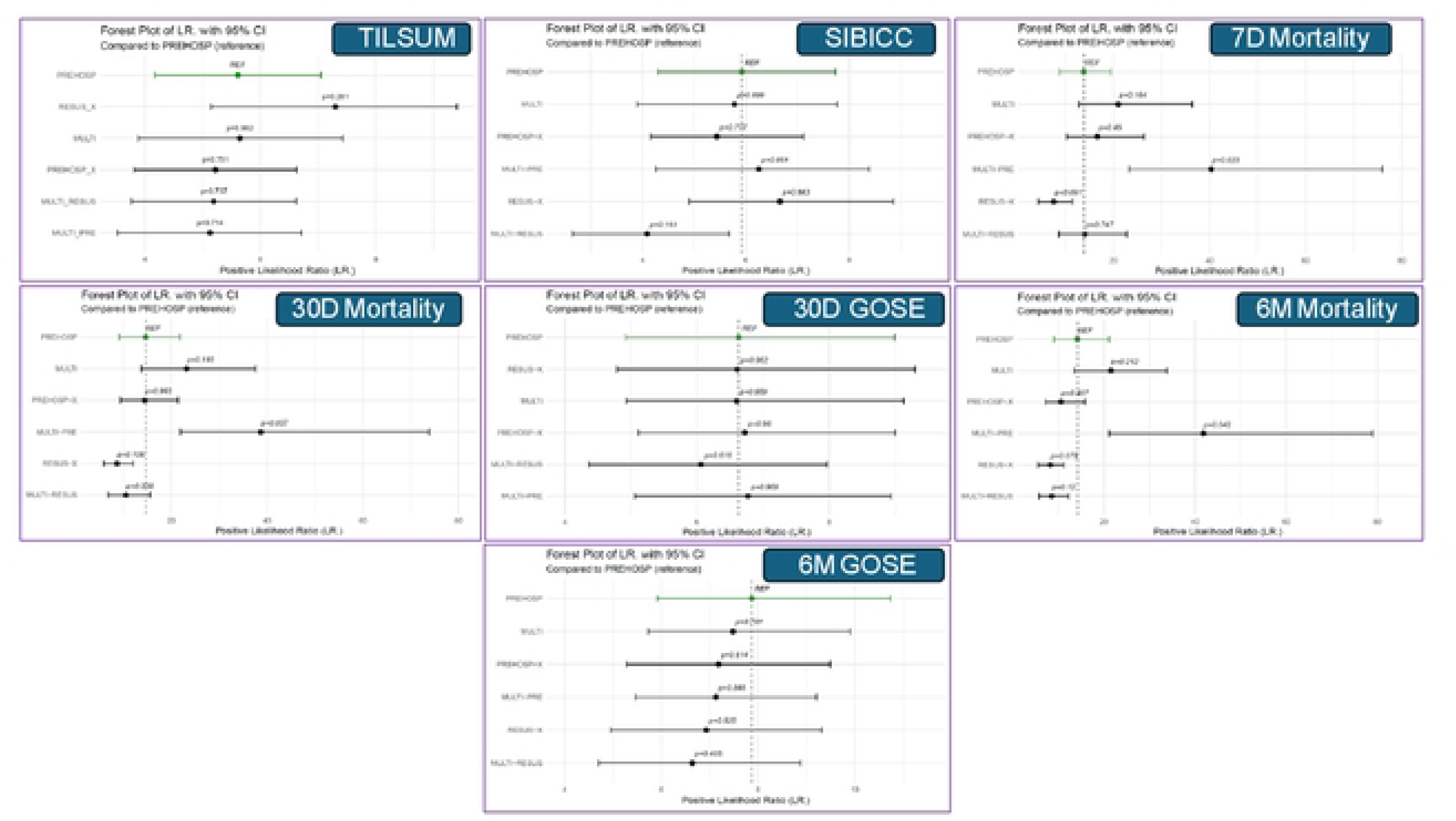
Forest Plot per outcome and for all seven models compared to PREHOSP as reference model. SIBICC= Seattle International Traumatic Brain Injury Consensus Conference. TILSUM=Therapeutic Intensity Level. D= day. M= Month. GOSE= Glasgow outcome scale extended

CT-TIQUA demonstrated significantly lower positive likelihood ratios than MULTI-PRE for 30-day (12.75, 95% CI 6.53-22.94 vs 38.70, 95% CI 21.93-73.79) and 6-month mortality (14.02, 95% CI 8.19-24.80 vs 41.83, 95% CI 21.08-78.95; see Figure 4).

**Figure 4.**
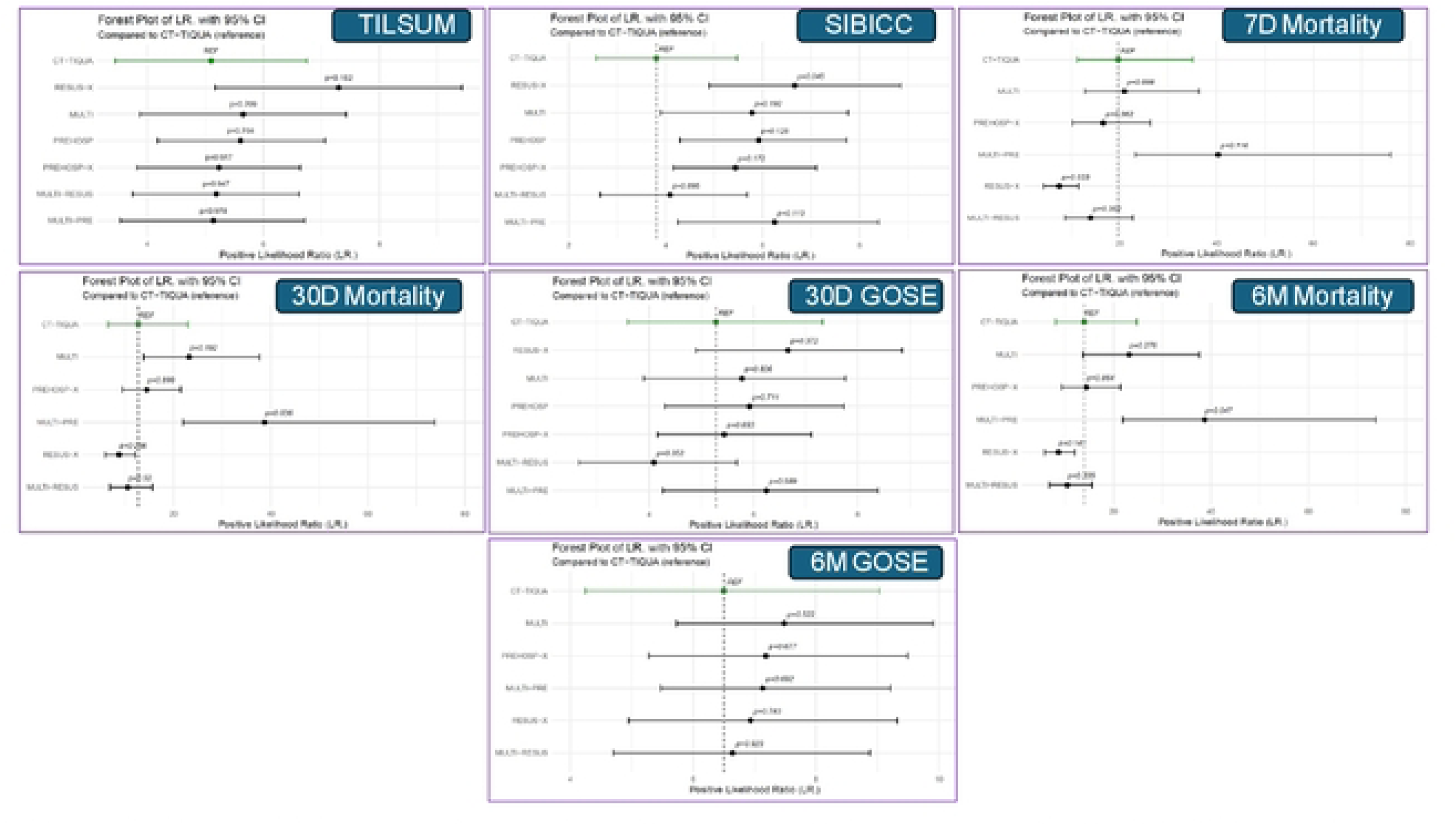
Forest Plot per outcome for all seven models compared to CT-TIQUA as reference model. SIBICC= Seattle International Traumatic Brain Injury Consensus Conference. TILSUM=Therapeutic Intensity Level. D= day. M= Month. GOSE= Glasgow outcome scale extended

### Secondary Outcomes

AUC-ROC across all models and outputs ranged from 0.80 (95% CI 0.69-0.90) for CT-TIQUA 6-month mortality to 0.97 (95% CI 0.95-0.98 and 0.93-0.99) for MULTI and MULTI-PRE 7-day mortality prediction. MULTI-PRE achieved the lowest Brier score of 0.03 (95% CI 0.02-0.04) for 30-day and 6-month mortality. No sex or age bias was identified.

Computation time for machine learning models required significantly less time for training and inference than deep learning models. As example, training of the model and inference of a single case of the Convolutional Neural Network CT-TIQUA compared to the Gradient Boost to predict 6 month GOSE lasted about 100 longer (Training: 390 versus 40046 seconds; inference 0.0034 versus 342 seconds) on a AMD EPYC 7443 24-Core Processor (see supporting information).

The training to predict 6 month GOSE for of the carbon Gradient Boost model required 1.3 of g CO₂eq compared to 377.7 g CO₂eq for the Convolutional Neural Network CT-TIQUA on the aforementioned processor for the energy mix in France. Inference for a single case of both models required 0.000058 (SD 0.000025) compared to mean=0.00059 (SD 0.00014) g CO₂eq, a ten-fold increase (see supporting information) Net clinical benefit at 10% decision threshold ranged from 0.029 for CT-TIQUA predicting SIBICC tier to 0.06 for MULTI predicting 30-day mortality. Net clinical benefit of 0.06 indicates 6 additional appropriate decisions per 100 patients versus not using the prediction tool. Benefits were comparable across models despite varying computational complexity (Figure 5).

**Figure 5.**
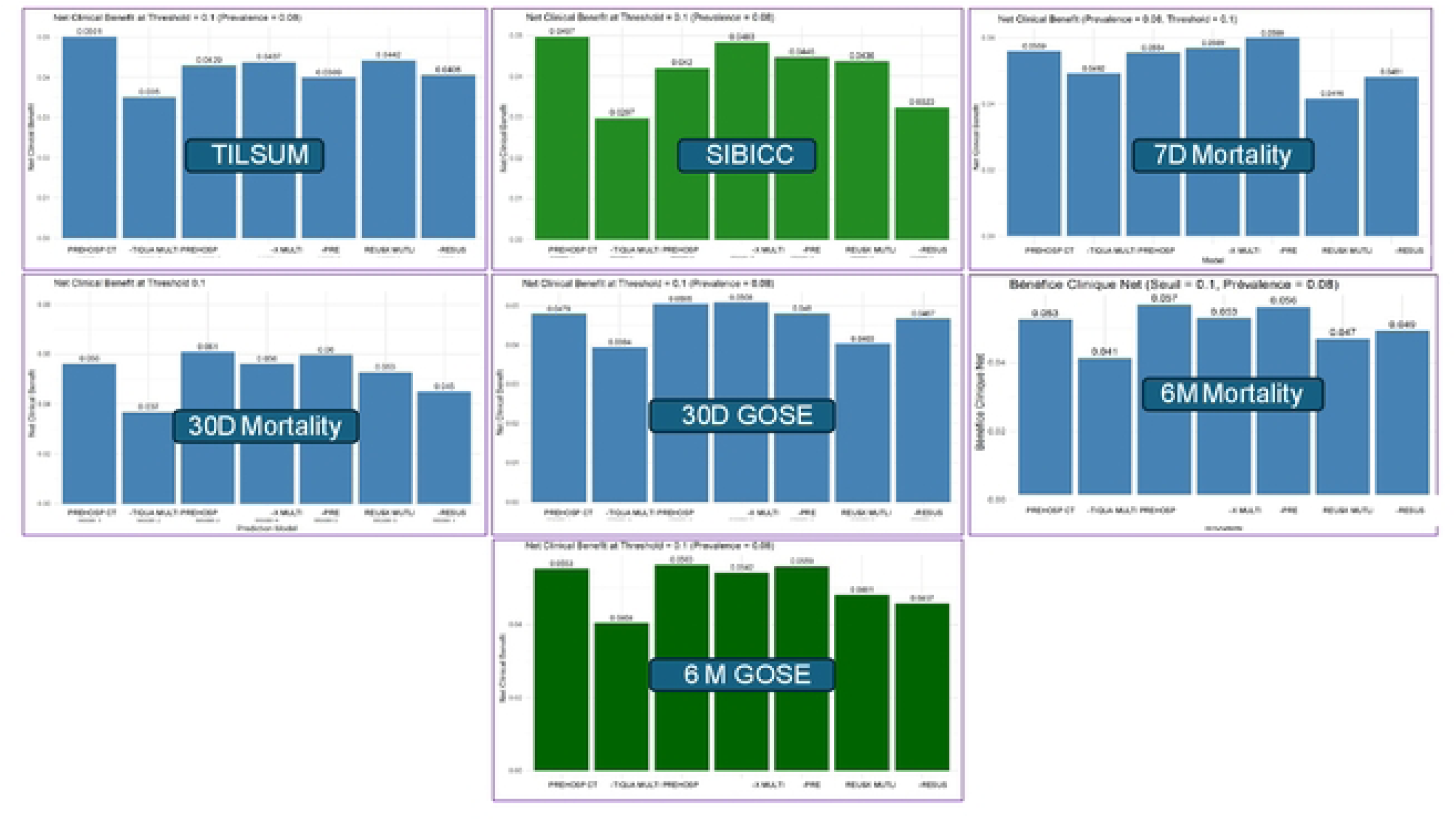
Net Clinical Benefit according to Vickers et al for a 10% decision threshold across all seven models. SIBICC= Seattle International Traumatic Brain Injury Consensus Conference. TILSUM=Therapeutic Intensity Level. D= day. M= Month. GOSE= Glasgow outcome scale extended

## DISCUSSION

The FASTDIAG I study provided external validation of PREHOSP and CT-TIQUA models and compared predictive performance of five multiparameter models to two pauci-parameter models for TBI pathway prediction. Multiparameter models did not demonstrate systematic superiority in predictive performance or net clinical benefit across all outputs. Combining CT segmentation by Convolutional Neural Networks with clinical information increased predictive performance but required substantially higher computational and informational resources and carbon foofprint for comparable net clinical benefit.

Outcome prediction in neurotrauma challenges even experienced clinicians, (20) yet they frequently must anticipate appropriate neurocritical care levels and make prognostic decisions. Excessive neurocritical care risks iatrogenesis or unfavourable long-term outcomes, while insufficient care increases secondary brain injury.(21) The ability to anticipate and quantify prognostic scenarios constitutes crucial elements for communication with families and care teams. These dilemmas increase cognitive load for providers and may delay neurocritical care initiation.

Several groups have deployed machine and deep learning approaches for TBI prediction in mostly retrospective studies. Malhotra et al reviewed TBI mortality and outcome prediction across 39 studies comprising 592,323 patients, finding AUC ranges from 0.68 to 0.95.(12) Pooled results across fourteen studies showed algorithms outperformed CRASH and IMPACT scores.(22) A systematic review of ICP increase prediction models across eleven studies observed AUCs ranging from 0.65 to 0.94 (average 0.85) with high overall bias risk.(3) Radiomic approaches show acceptable performance for detecting and quantifying intracranial injuries.(23) To our knowledge, no systematic review compares radiomic approaches for outcome prediction. To our knowledge, no systematic review is available comparing radiomic approaches to predict patient outcome. As individual high evidence study, Monteiro el. validated a convolutional neural network to predict different types of TBI injuries with AUC ranging from 0.8 to 0.96 without predicting patient outcome.(24) HajiEsmailPoor et al. compared radiomic models for ICH for outcome prediction encompassing TBI and showed best AUC of 0.83.(25) Only few studies attempted to predict actionable patient needs such as neurosurgery or neurocritical care level on the patient pathway and no systematic review is available for comparison. (10,11,26,27)

Few models attempt predictions along the TBI patient pathway combining multiple outputs and input variables. Most studies do not report the complete spectrum of recommended metrics, with AUC remaining most frequently reported. Machine learning tools generally outperform simple regression.³ Net clinical benefit, decision curves, and technological readiness are rarely reported. The predictive-efficiency trade-off or computational return on investment are rarely considered.⁵,⁹

The present study compares favourably to aforementioned studies across all models and outputs. For example, Courville et al observed AUCs from 0.78 to 0.89 for 6-month Glasgow Outcome Scale Extended prediction compared with FASTDIAG models ranging from 0.80 (CT-TIQUA) to 0.96 (PREHOSP and MULTI).³ The FASTDIAG study contributes novel aspects. First, it demonstrates feasibility of predicting the clinical TBI pathway from short-term actionable needs (neurocritical care level) to long-term outcomes (6-month Glasgow Outcome Scale Extended) with two pauci-parameter and five multiparameter models. Second, results suggest comparable clinical benefit between pauci-parameter and multiparameter models, particularly PREHOSP. Standalone CT-TIQUA performance confirms clinical intuition that CT analysis alone, including radiomic approaches, requires clinical context interpretation. Models containing prehospital features showed more consistent performance than those with resuscitation room features, especially when combined with CT segmentation. Third, despite better predictive performance of multiparameter MULTI and MULTI-PRE algorithms combining clinical data and radiomic features, clinical benefit was not overwhelming and came at significantly higher informational and computational cost and higher carbon footprint.

The deep learning models generated a 10 to 100 fold higher energy consumption and carbon footprint for comparable clinical performance as measured in net clinical benefit. In comparison the training of the convolutional neural network model generated six times more CO2 than a standard single head CT-scan for the energy-mix in France and the equivalent of a CT-head in the US or Germany (see supporting information).

This observation is important since many published models rely on high-quality, multiparameter data from complex electronic health record ecosystems.(8,28,29) These informatic ecosystems are not widely available, even in high-income countries, due to cost, lack of expertise, and regulatory barriers.(30) This explains variability in data extraction methods, missing values, and data quality across institutions, affecting generalizability and external validation.(9) Different imaging protocols or electronic health record standards can significantly affect trained model performance when deployed in diverse settings or lead to overfitting, especially for complex models like deep neural networks with limited training data. Most published algorithms were developed in high socioeconomic index countries, yet severe TBI burden is highest in low socioeconomic index countries.(31) Workable enhanced decision support requires simple, reliable algorithms.

The present study highlights the need to assess predictive-efficiency trade-offs such as computational needs and ecological impact and resource consumption and compare systematically to predictive performance and clinical benefit for enhanced decision support tools. This addresses the crucial question: what informational and predictive gain does this model provide at what computational, informational, and deployment and environmental cost? This question is crucial to close the persistent knowledge gap between model development, external validation, deployment, patient benefit assessment, and comparison against human clinicians in real clinical workflows.(5) To our knowledge, this is the first validation study to address these issues and the one of the first to compare the carbon footprint of clinical prediction models to their performance in particular in the context of critical care. (7)

### Limitations

Several limitations warrant consideration. First, the retrospective, monocentric design may introduce center bias. Investigators attempted to reduce confirmation bias by having data retrieval performed by clinicians uninvolved in model training and validation. However, automatic electronic health record data extraction is also error-prone and requires resource-intensive human verification. Second, the dataset is small compared with other studies, and small datasets systematically overestimate predictive performance. Notably, this study externally validates previously published PREHOSP and CT-TIQUA models, with consistent performance across development, validation, and external validation phases. Third, the included feature list may seem limited, and additional features such as biomarkers or physiological variables like ICP could improve predictive performance. However, investigators focused on widely available routine features from the initial phase to increase feasibility, data availability, external validity, and operationality. ICP and ICP dose, being surrogate markers of neurocritical care intensity, were excluded to reduce possible collinearity and collider bias. Finally, bias assessment was performed only for age and sex, as the dataset lacked socioeconomic data and French legislation prevents ethnic data collection.

## CONCLUSIONS

The FASTDIAG I study provided external validation of two pauci-parameter models and compared their predictive performance with five multiparameter models for TBI patient pathway prediction. Multiparameter models did not demonstrate systematic superiority in performance or net clinical benefit across all pathway outputs. Combining CT segmentation by Convolutional Neural Networks with clinical information increased predictive performance but generated higher computational, informational and ecological costs for comparable net clinical benefit. These findings highlight the importance of systematically considering predictive-efficiency trade-offs to facilitate model deployment and patient benefit assessment compared to human clinicians in real clinical workflows.

## METHODS

### Study Design and Setting

This retrospective single-center comparative and external validation study was conducted at Grenoble University Hospital (GUH), a level 1 trauma center with dedicated neurocritical care. Patients were managed according to standard operating procedures based on national and international guidelines. The institutional review board of Grenoble University hospital approved the protocol on the 22/11/2024 (File nr 38RC23.0351, CHU Grenoble, Grenoble, France). All patients or their next of kin received written and formal notification with the opt-out possibility to opt-out of the study; four patients decided to opt out. The manuscript follows AI APPRAISE TOOL guidelines (12).

### Patient Selection

All adults admitted to the trauma resuscitation unit between August 1, 2020, and December 31, 2021, were screened. Inclusion criteria comprised TBI defined as Glasgow Coma Scale <9 and/or documented intracranial injuries on admission CT scan. Data were retrieved by two expert clinicians blinded to model outputs. CT scans were retrieved from hospital imaging systems and stored anonymously. Observations with missing predictor variables or outcomes were excluded without imputation. The corresponding CT scan was retrieved from the hospital image processing system and corresponding ID assigned. The anonymized CT scan with corresponding ID was stored on a safe imaging platform (*shanoir*) in DICOM format. All data (clinical and images) were stored on a safe server after anonymization and ID number assignment and data management performed centrally by professionals blinded to the model output. Figure 1 summarizes the flowchart and data processing.

### Prediction Models

Pauci-Parameter Models

PREHOSP uses 15 routinely collected prehospital predictors (Glasgow Coma Scale components, age, capillary haemoglobin, blood pressure, heart rate, pupil anomaly, limb ischemia, pelvic injury, cardiac arrest, external bleeding, penetrating trauma, vasopressor use). This classification model employs light Gradient Boosting,(13) initially trained to predict emergency neurosurgery within 24 hours and need for ICP monitoring (supporting information 2).

CT-TIQUA analyses admission CT scans performed within 60 minutes of resuscitation room admission. The model performs four steps: extraction (removes bone), registration (locates lesions topographically), segmentation (identifies injury types using deep learning), and quantification (computes volumes). CT-TIQUA recognizes seven distinct TBI classes using Convolutional Neural Networks: intraparenchymal, subdural, epidural hematoma, intraventricular and subarachnoid hemorrhage, petechiae, and edema (supporting information 3). The initial CT-TIQUA model was improved, first by testing three different brain extraction methods; the TOTALSEGMENTATOR (14) approach proved to be the best performing with an AUC of 0.98 tested against manual ground truth brain masks. Second, three different *atlas registration* techniques were explored; a combination of Histogram matching and Advanced Normalization Tools appeared to be the most reliable.(15) The supporting information provides details on the development process.

### Multiparameter Models

Five multiparameter models were developed: MULTI (combining PREHOSP and CT-TIQUA features), PREHOSP-X (relying on all available 23 prehospital variables), RESUS-X (relying on all available 18 resuscitation room variables), MULTI-PRE (PREHOSP-X combined with CT-TIQUA), and MULTI-RESUS (RESUS-X combined with CT-TIQUA). PREHOSP-X and RESUS-X employed light Gradient Boosting (supporting information 4).

In summary, seven models were available. Two pauci-parameter models PREHOSP and CT-TIQUA and five multi-parameter models, MULTI, PREHOSP-X, RESUS-X and MULTI-PRE and MULTI-RESUS.

### Model Outputs

All seven models predicted (Figure 2):

1. Highest neurocritical care intensity within seven days per TILSUM classification (dichotomized as >level 0); http://www.tbi-impact.org/cde/mod_templates/T_TIL.9.1.pdf)(16)
2. Highest neurocritical care per Seattle International Severe TBI Consensus Conference (SIBICC)(17) (dichotomized as >tier 0)
3. Mortality at 7-day, 30-day, and 6-months
4. Extended Glasgow Outcome Scale at 30-day and 6-months

Each prediction constituted binary classification. Models underwent hyperparameter optimization, class imbalance correction, and cross-validation (see supporting information for details).

### Primary assessment criterion

The primary assessment criterion was the positive Likelihood ratio with 95% confidence interval. All other metrics were considered as secondary outcome. The primary analysis concerned the comparison between the positive likelihood ratios between the pauci-parameter models PREHOSP and CT-TIQUA and all multi-parameter models (MULTI, PREHOSP-X, RESUS-X and MULTI-PRE and MULTI-RESUS) for all seven outputs (see above). All other metrics, execution time and the additional analysis recruiting all available prehospital and resuscitation room variables (preceding paragraph) were considered as secondary outcome criteria.

### Sample size calculation

The sample size was determined based on the comparison of the light Gradient Boosting prediction and the truly observed rate of the composite outcome criterion *need for neurosurgery and ICP probe placement*. The prevalence of the outcome in a pilot study was 15%, sensitivity 84% and specificity of 87% for the light Gradient Boosting based on a pilot study.(10) With a type 1 error of 5%, a prevalence of the main output of 15% and to obtain a positive likelihood ratio around 6 for 95% confidence interval with a width of 0.255, a sample of 550 patients was necessary. (18)

### Computation efficiency and Carbon Footprint calculation

Computation time for training and single case inference for the Machine Learning and Deep learning (radiomics) models were expressed in seconds. The respective carbon footprint were calculated from development to application, The method used has been described at https://codecarbon.io/. In summary, the following formula is used:

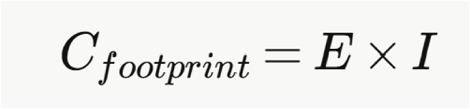

Where *Cfootprint* is carbon footprint in kg of CO₂ equivalent (CO₂eq), *E*= energy consumed by the computation in kilowatt-hours (kWh), *I*= carbon intensity of electricity in the geographic location of the hardware, in kg CO₂eq per kWh. CodeCarbon computes the carbon footprint of your code by measuring at fixed intervals, e.g. every 15 seconds, the electricity consumption of the GPU, CPU and RAM on which the code is executed. The package also monitors the duration of code execution and uses this information to compute the total electricity consumption. CodeCarbon retrieves information about the carbon intensity of the electricity in the geographic location of the hardware. The calculation of the carbon footprint included the development and training phase of each model.

### Data analysis

All data were analyzed by a data engineer and clinician researcher (TFD, AK) supervised by a senior clinician researcher (TG) and two research directors (BL, JJ). Continuous data were described as median (quartiles 1–3) and categorical variables as count (percentages).

The true observed rate of all listed outputs in the patient cohort was considered as reference. The individual (pauci-parameter) and combined (multi-parameter) model performance was assessed using the following metrics: AUC ROC, sensitivity, specificity, positive, negative predictive value, Brier score, Youden Index, F2, accuracy and precision and the positive and negative likelihood ratio (LR+ and LR-). The positive likelihood ratio of the seven models considered for each model and their combination were compared through a z-score with a 95% confidence interval computed by bootstrapping, estimate the standard error from the CI and apply pairwise z-tests on log(LR+) values with Bonferroni correction for multiple comparisons. The net clinical benefit for each prediction model and combined use was calculated according to Vickers et al with a decision threshold of 10%. (19) Potential bias of age and sex were compared for all metrics. All calculations and developments were performed on Python (version 3.11.0) or R and R studio (version 2024.12.1).

## ARTICLE INFORMATION

**Ethical Approval and Consent to participate:** The institutional review board of Grenoble University hospital approved the protocol on the 22/11/2024. All patients or their next of kin received written and formal notification with the opt-out possibility to opt-out of the study; four patients decided to opted out.

**Consent for publication:** all authors consented to the publication and the manuscript has not been published elsewhere and is not under consideration by another journal. No artificial intelligence was used to write this manuscript

**Compliance with Checklist:** AI APPRAISE guideline

**Availability of supporting data:** Anonymized data collected for this study will be made available on request from the corresponding author. The computer programs used to train each model and compute metrics with their confidence intervals is available on GitHub https://github.com/nifm-gin/FastDiag.

No generative AI was used to conceive or write or modify the manuscript.

## Authors’ contributions

Design, Data analysis, model construction, writing of manuscript: T Gauss (also data collection), T Fehr-Delude, A Kalimouttou, B Lemasson Model construction: G Brelurut, S Medjkoune, C Brossard Data collection, critical review: O Seddiki, C Sanchez Critical review: JD Moyer, J Greze, A Lazard, A Krainik, T Boulier, K Lagarde, P Bouzat

## Data Availability

Anonymized data collected for this study will be made available on request from the corresponding author. The computer programs used to train each model and compute metrics with their confidence intervals is available on GitHub https://github.com/nifm-gin/FastDiag. No generative AI was used to conceive or write or modify the manuscript.

## Acknowledgements

not relevant

## Supporting information

1) AI APPRAISE tool

2) Initial PREHOSP, neurosurgery prediction model

3) CT scan radiomic analysis, CT-TIQUA

4) List of variables

5) Combined prediction models

6) Shapley values, false positive and negative rates, example PREHOSP

7) Combined model metrics

8) Decision Curve Analysis

9) TILSUM class

10) SIBICC classification

11) Carbon footprint analysis

## REFERENCES

1. Korley FK, Peacock WF, Eckner JT, Maio R, Levin S, Bechtold KT, et al. Clinical Gestalt for Early Prediction of Delayed Functional and Symptomatic Recovery From Mild Traumatic Brain Injury Is Inadequate. Panagos P, editor. Acad Emerg Med. 2019 Dec;26(12):1384–7. doi:10.1111/acem.13844

2. Courville E, Kazim SF, Vellek J, Tarawneh O, Stack J, Roster K, et al. Machine learning algorithms for predicting outcomes of traumatic brain injury: A systematic review and meta-analysis. Surg Neurol Int. 2023 Jul 28;14:262. doi:10.25259/sni_312_2023

3. van Hal ST, van der Jagt M, van Genderen ME, Gommers D, Veenland JF. Using Artificial Intelligence to Predict Intracranial Hypertension in Patients After Traumatic Brain Injury: A Systematic Review. Neurocrit Care. 2024 Aug;41(1):285–96. doi:10.1007/s12028-023-01910-2 PubMed PMID: 38212559; PubMed Central PMCID: PMC11335950.

4. Fong N, Feng J, Hubbard A, Dang LE, Pirracchio R. IntraCranial pressure prediction AlgoRithm using machinE learning (I-CARE): Training and Validation Study. Crit Care Explor. 2023 Dec 28;6(1):e1024. doi:10.1097/cce.0000000000001024

5. Berkhout WEM, Van Wijngaarden JJ, Workum JD, Van De Sande D, Hilling DE, Jung C, et al. Operationalization of Artificial Intelligence Applications in the Intensive Care Unit: A Systematic Review. JAMA Netw Open. 2025 Jul 23;8(7):e2522866. doi:10.1001/jamanetworkopen.2025.22866

6. Gauss T, Moyer JD, Colas C, Pichon M, Delhaye N, Werner M, et al. Pilot deployment of a machine-learning enhanced prediction of need for hemorrhage resuscitation after trauma - the ShockMatrix pilot study. BMC Med Inform Decis Mak. 2024 Oct 28;24(1):315. doi:10.1186/s12911-024-02723-9 PubMed PMID: 39468585; PubMed Central PMCID: PMC11520814.

7. Truhn D, Müller-Franzes G, Kather JN. The ecological footprint of medical AI. Eur Radiol. 2023 Aug 15;34(2):1176–8. doi:10.1007/s00330-023-10123-2

8. Pinsky M, Dubrawski A, Clermont G. Intelligent Clinical Decision Support. Sensors. 2022 Feb 12;22(4):1408. doi:10.3390/s22041408

9. Ellis RJ, Sander RM, Limon A. Twelve key challenges in medical machine learning and solutions. Intell-Based Med. 2022;6:100068. doi:10.1016/j.ibmed.2022.100068

10. Moyer JD, Lee P, Bernard C, Henry L, Lang E, Cook F, et al. Machine learning-based prediction of emergency neurosurgery within 24 h after moderate to severe traumatic brain injury. World J Emerg Surg WJES. 2022 Aug 3;17(1):42. doi:10.1186/s13017-022-00449-5 PubMed PMID: 35922831; PubMed Central PMCID: PMC9351267.

11. Brossard C, Grèze J, de Busschère JA, Attyé A, Richard M, Tornior FD, et al. Prediction of therapeutic intensity level from automatic multiclass segmentation of traumatic brain injury lesions on CT-scans. Sci Rep. 2023 Nov 17;13(1):20155. doi:10.1038/s41598-023-46945-9 PubMed PMID: 37978266; PubMed Central PMCID: PMC10656472.

12. Kwong JCC, Khondker A, Lajkosz K, McDermott MBA, Frigola XB, McCradden MD, et al. APPRAISE-AI Tool for Quantitative Evaluation of AI Studies for Clinical Decision Support. JAMA Netw Open. 2023 Sep 25;6(9):e2335377. doi:10.1001/jamanetworkopen.2023.35377

13. Pedregosa F, Varoquaux G, Gramfort A, Michel V, Thirion B, Grisel O, et al. Scikit-learn: Machine Learning in Python [Internet]. 2012. doi:10.48550/ARXIV.1201.0490

14. Li Y, Wynne JF, Wu Y, Qiu RLJ, Tian S, Wang T, et al. Automatic medical imaging segmentation via self-supervising large-scale convolutional neural networks. Radiother Oncol. 2025 Mar;204:110711. doi:10.1016/j.radonc.2025.110711

15. Avants B, Epstein C, Grossman M, Gee J. Symmetric diffeomorphic image registration with cross-correlation: Evaluating automated labeling of elderly and neurodegenerative brain. Med Image Anal. 2008 Feb;12(1):26–41. doi:10.1016/j.media.2007.06.004

16. Bhattacharyay S, Beqiri E, Zuercher P, Wilson L, Steyerberg EW, Nelson DW, et al. Therapy Intensity Level Scale for Traumatic Brain Injury: Clinimetric Assessment on Neuro-Monitored Patients Across 52 European Intensive Care Units. J Neurotrauma. 2024 Apr 1;41(7–8):887–909. doi:10.1089/neu.2023.0377

17. Hawryluk GWJ, Aguilera S, Buki A, Bulger E, Citerio G, Cooper DJ, et al. A management algorithm for patients with intracranial pressure monitoring: the Seattle International Severe Traumatic Brain Injury Consensus Conference (SIBICC). Intensive Care Med. 2019 Dec;45(12):1783–94. doi:10.1007/s00134-019-05805-9 PubMed PMID: 31659383; PubMed Central PMCID: PMC6863785.

18. Bachmann LM, Puhan MA, Riet GT, Bossuyt PM. Sample sizes of studies on diagnostic accuracy: literature survey. BMJ. 2006 May 13;332(7550):1127–9. doi:10.1136/bmj.38793.637789.2F

19. Vickers AJ, Elkin EB. Decision Curve Analysis: A Novel Method for Evaluating Prediction Models. Med Decis Making. 2006 Nov;26(6):565–74. doi:10.1177/0272989X06295361

20. Bonds B, Dhanda A, Wade C, Diaz C, Massetti J, Stein DM. Prognostication of Mortality and Long-Term Functional Outcomes Following Traumatic Brain Injury: Can We Do Better? J Neurotrauma. 2021 Apr 15;38(8):1168–76. doi:10.1089/neu.2014.3742

21. Robba C, McCredie V, Chesnut RM, Citerio G, Gauss T, Hawryluk GWJ, et al. Traumatic brain injury management in the intensive care unit: standard of care and knowledge gaps. Intensive Care Med. 2025 Jun;51(6):1112–27. doi:10.1007/s00134-025-07967-1

22. Malhotra AK, Shakil H, Smith CW, Huang YQ, Kwong JCC, Thorpe KE, et al. Predicting outcomes after moderate and severe traumatic brain injury using artificial intelligence: a systematic review. Npj Digit Med. 2025 Jun 18;8(1):373. doi:10.1038/s41746-025-01714-y

23. Hibi A, Jaberipour M, Cusimano MD, Bilbily A, Krishnan RG, Aviv RI, et al. Automated identification and quantification of traumatic brain injury from CT scans: Are we there yet? Medicine (Baltimore). 2022 Nov 25;101(47):e31848. doi:10.1097/MD.0000000000031848

24. Monteiro M, Newcombe VFJ, Mathieu F, Adatia K, Kamnitsas K, Ferrante E, et al. Multiclass semantic segmentation and quantification of traumatic brain injury lesions on head CT using deep learning: an algorithm development and multicentre validation study. Lancet Digit Health. 2020 Jun;2(6):e314–22. doi:10.1016/S2589-7500(20)30085-6

25. HajiEsmailPoor Z, Kargar Z, Baradaran M, Shojaeshafiei F, Tabnak P, Mandalou L, et al. Prognostic value of CT scan-based radiomics in intracerebral hemorrhage patients: A systematic review and meta-analysis. Eur J Radiol. 2024 Sep;178:111652. doi:10.1016/j.ejrad.2024.111652

26. Zhu G, Ozkara BB, Chen H, Zhou B, Jiang B, Ding VY, et al. Enhancing hospital course and outcome prediction in patients with traumatic brain injury: A machine learning study. Neuroradiol J. 2024 Feb;37(1):74–83. doi:10.1177/19714009231212364 PubMed PMID: 37921691; PubMed Central PMCID: PMC10863571.

27. Habibzadeh A, Khademolhosseini S, Kouhpayeh A, Niakan A, Asadi MA, Ghasemi H, et al. Machine learning-based models to predict the need for neurosurgical intervention after moderate traumatic brain injury. Health Sci Rep. 2023 Nov;6(11). doi:10.1002/hsr2.1666

28. Khaled A, Sabir M, Qureshi R, Caruso CM, Guarrasi V, Xiang S, et al. Leveraging MIMIC Datasets for Better Digital Health: A Review on Open Problems, Progress Highlights, and Future Promises [Internet]. arXiv; 2025 [cited 2025 Aug 10]. Available from: https://arxiv.org/abs/2506.12808doi:10.48550/ARXIV.2506.12808

29. Weiskopf NG, Bakken S, Hripcsak G, Weng C. A Data Quality Assessment Guideline for Electronic Health Record Data Reuse. EGEMs Gener Evid Methods Improve Patient Outcomes. 2017 Sep 4;5(1):14. doi:10.5334/egems.218

30. Gauss T, Bouzat P, Josse J. From bedside to data and back to bedside: why do we need better data to perform trauma phenotyping and improve trauma care? Intensive Care Med. 2025 Jun;51(6):1161–3. doi:10.1007/s00134-025-07946-6

31. Dewan MC, Rattani A, Gupta S, Baticulon RE, Hung YC, Punchak M, et al. Estimating the global incidence of traumatic brain injury. J Neurosurg. 2019 Apr 1;130(4):1080–97. doi:10.3171/2017.10.JNS17352 PubMed PMID: 29701556.

